# Accurate prediction of neurologic changes in critically ill infants using pose AI

**DOI:** 10.1101/2024.04.17.24305953

**Authors:** Alec Gleason, Florian Richter, Nathalia Beller, Naveen Arivazhagan, Rui Feng, Emma Holmes, Benjamin S Glicksberg, Sarah U Morton, Maite La Vega-Talbott, Madeline Fields, Katherine Guttmann, Girish N Nadkarni, Felix Richter

## Abstract

Infant alertness and neurologic changes can reflect life-threatening pathology but are assessed by exam, which can be intermittent and subjective. Reliable, continuous methods are needed. We hypothesized that our computer vision method to track movement, pose AI, could predict neurologic changes in the neonatal intensive care unit (NICU). We collected 4,705 hours of video linked to electroencephalograms (EEG) from 115 infants. We trained a deep learning pose algorithm that accurately predicted anatomic landmarks in three evaluation sets (ROC-AUCs 0.83–0.94), showing feasibility of applying pose AI in an ICU. We then trained classifiers on landmarks from pose AI and observed high performance for sedation (ROC-AUCs 0.87–0.91) and cerebral dysfunction (ROC-AUCs 0.76–0.91), demonstrating that an EEG diagnosis can be predicted from video data alone. Taken together, deep learning with pose AI may offer a scalable, minimally invasive method for neuro-telemetry in the NICU.

## INTRODUCTION

Infant alertness is considered the most sensitive piece of the neurologic exam, reflecting integrity throughout the central nervous system.^1,2^ Infant mental status changes can be due to encephalopathy, sedation, or other causes and are highly dynamic, necessitating continuous assessment. Encephalopathy can be caused by a variety of diagnoses which require rapid identification and treatment (*e.g.*, hypoxic, metabolic, and infectious etiologies).^3^ Lethargy, a sign of encephalopathy, is one of the most important indicators of neonatal sepsis.^4^ While encephalopathy is an example of pathology that impacts alertness, we also purposefully manipulate infant mental status, most commonly with sedative medications. Titrating the appropriate level of sedation is challenging in any patient population, but more so in infants due to their inability to communicate and the extreme pharmacokinetic variability of sedative medications. In infants, these medications have longer half-lives, high rates of tachyphylaxis, opioid antagonist metabolites, and more pronounced respiratory depression.^5,6^ Altogether, we routinely evaluate mental status to make life-saving decisions in the neonatal intensive care unit (NICU).

Despite its importance, our ability to continuously quantify and monitor infant alertness is limited. The physical exam is commonly employed, but provides a single snapshot, is subjective, and can be delayed.^1,2^ Validated instruments, such as the N- PASS for sedation^7^ or modified Sarnat for encephalopathy,^8,9^ mitigate exam subjectivity with high inter-rater reliability^10^ but are labor-intensive and not continuous.

Electroencephalography (EEG) is a continuous record of neurologic activity. However, its implementation requires specialized staff and equipment not available at every NICU,^11^ and it carries the risk of pressure injuries with prolonged use.^12^ Applicability of EEG to alertness is also limited, as there is no association between EEG and neonatal depth of sedation.^13^ Furthermore, although continuous EEG can assess encephalopathy by indicating the degree of cerebral dysfunction, there is limited clinical adoption aside from evaluation for therapeutic hypothermia. This is likely owing to EEG’s high inter-rater variability and challenging interpretability among bedside providers.^14,15^ Overall, determining the level of an infant’s alertness remains a challenge, despite being fundamental to clinical care in the NICU.

One scalable approach to address this unmet need is to augment the physical exam using computer vision. We hypothesized that pose artificial intelligence (AI),^16,17^ a deep learning approach to track anatomic landmarks, could be used to continuously characterize changes in neurological phenotypes in the NICU. In this study, we successfully applied pose AI to a large dataset (4,705 video-EEG hours, 115 infants) and used pose to accurately predict sedation and cerebral dysfunction in critically ill infants (**Figure 1**).

**Figure 1.**
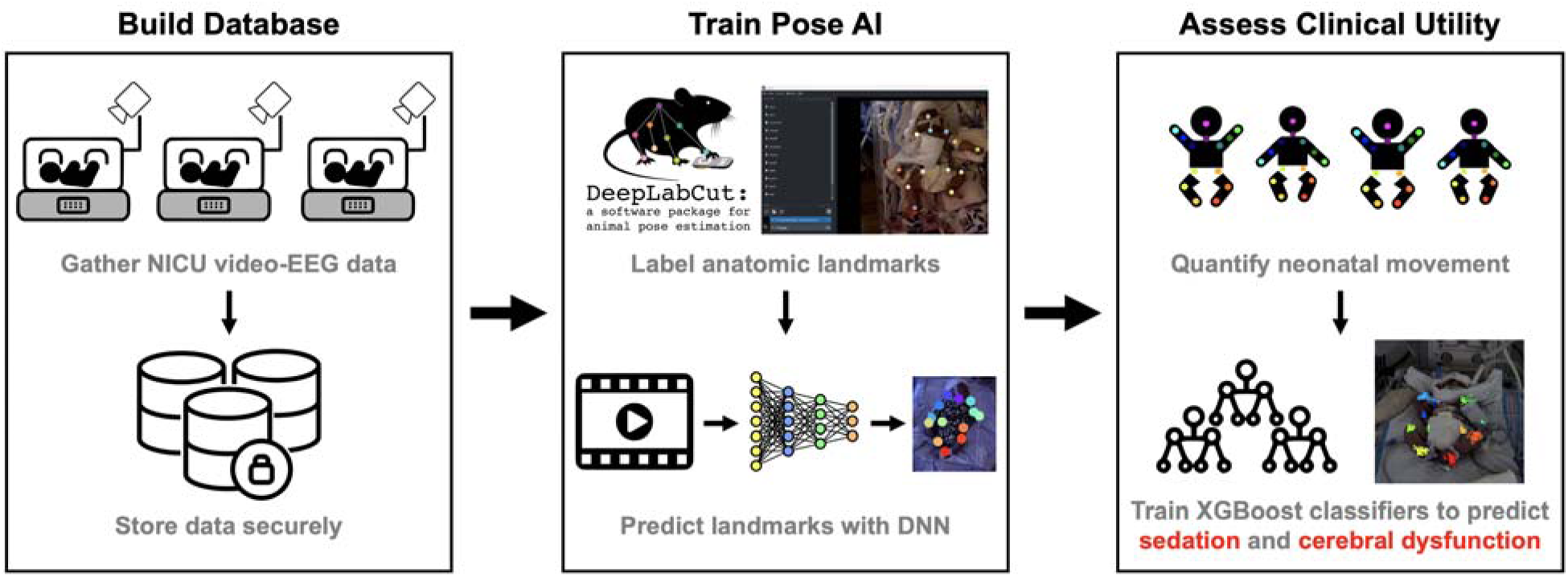
Application of Pose AI to critically ill infants. We built a large database of video-EEG data (N=115 infants, 4,705 hours of video, 10.4 Tb) that is stored on a HIPAA-compliant supercomputing cluster. We then trained/tested a Pose AI algorithm on 2712 manually labelled video frames with DeepLabCut to predict infant anatomic landmarks. Finally, we demonstrate clinical utility of quantifying movement and use infant movement features to predict sedation and cerebral dysfunction. DNN=deep neural network.

## RESULTS

### Cohort characteristics

Video-EEG and clinical data were collected from Feb 2021 to Dec 2022, during which data from 115 individuals met inclusion criteria (**Table 1**). Caregiver-reported race and ethnicity were obtained retrospectively from the electronic health record. Consistent with the routine use of video-EEG in therapeutic hypothermia, most infants were <1 month old (67%) and the most common underlying suspected/known pathology was hypoxic ischemic encephalopathy (21%). Both levetiracetam and phenobarbital were the most common first-line ASMs, possibly reflecting the recent publication of the NEOLEV2 trial.^34^

**Table 1.** Clinical characteristics of all patients included in this study.

| Descriptive factor |  | All (N=115) |
| --- | --- | --- |
| Gestational Age at Birth, *median (range)* |  | 38w1d<br>(23w0d – 41w1d) |
| Age in months, *N (%)* | < 1 | 77 (67.0) |
|  | [1 – 3) | 21 (18.3) |
|  | [3 – 6) | 10 (8.7) |
|  | ≥ 6 | 7 (6.1) |
| Sex, *N (%)* | Female | 53 (46.1) |
|  | Male | 62 (53.9) |
| Race, *N (%)* | American Indian or Alaskan | 5 (4.3) |
|  | Asian | 9 (7.8) |
|  | Black or African American | 27 (23.5) |
|  | Native Hawaiian or Pacific Islander | 1 (0.9) |
|  | White | 32 (27.8) |
|  | Other | 33 (28.7) |
|  | Unknown | 8 (7.0) |
| Ethnicity, *N (%)* | Hispanic/Latino | 33 (28.7) |
|  | Non-Hispanic | 58 (50.4) |
|  | Unknown | 24 (20.9) |
| Neurologic pathology, *N (%)* | Hypoxic ischemic encephalopathy | 24 (20.9) |
|  | Genetic or idiopathic epilepsy | 14 (12.2) |
|  | Intraventricular hemorrhage | 7 (6.1) |
|  | Stroke | 7 (6.1) |
|  | Structural brain malformation | 4 (3.5) |
|  | Other | 15 (13.0) |
|  | None | 44 (38.3) |
| CNS active medications*, *N (%)* | Levetiracetam | 5 (4.3) |
|  | Levetiracetam, phenobarbital | 9 (7.8) |
|  | Levetiracetam, phenobarbital, fos/phenytoin | 4 (3.5) |
|  | Phenobarbital | 16 (13.9) |
|  | Sedative infusion and any ASMs<sup>†</sup> | 12 (10.4) |
|  | Other combination of ASMs<sup>††</sup> | 5 (4.3) |
|  | None | 64 (55.7) |

\*All medications acting on the central nervous system (CNS) that were used at any point while the patient was undergoing the video-EEG study.
†ASMs=antiseizure medications
††Other ASMs were lacosamide, oxcarbazepine, topiramate, and vigabatrin

### Training and evaluating an infant pose recognition algorithm

We trained a computer vision algorithm, DeepLabCut, to predict infant pose. We trained DeepLabCut on 2,351 manually labeled frames from 99 infants, and evaluated performance within this training set, a set of held-out frames from the infants used for training, and a set of infants not used for training.^26,27^ We observed a low L2 pixel error (median L2_train_=3.2, L2_test-new-frames_=3.5, L2_test-new-infants_=4.6 pixels) comparable to prior work.^16^ This was consistent across all anatomic landmarks (**Figure 2a**). Further illustrating excellent model performance, we observed high ROC-AUCs >0.83 for landmark occlusion (**Figure 2b**) and that ≥95% of predictions were within a reference distance threshold, half the size of the head (**Figure 2c**). Finally, exemplary video frames during a seizure in a neonate with KCNQ2-related epilepsy syndrome showed predicted landmarks (colored) overlapping expected locations (**Figure 2d**). Having successfully trained a pose recognition algorithm on infants, we generated pose X and Y coordinates for all video data from all patients (N=4,705 hours from N=115 infants). This includes videos from six additional infants not initially available for training or evaluation, bringing the total from 109 to 115 infants for all downstream analyses.

**Figure 2.**
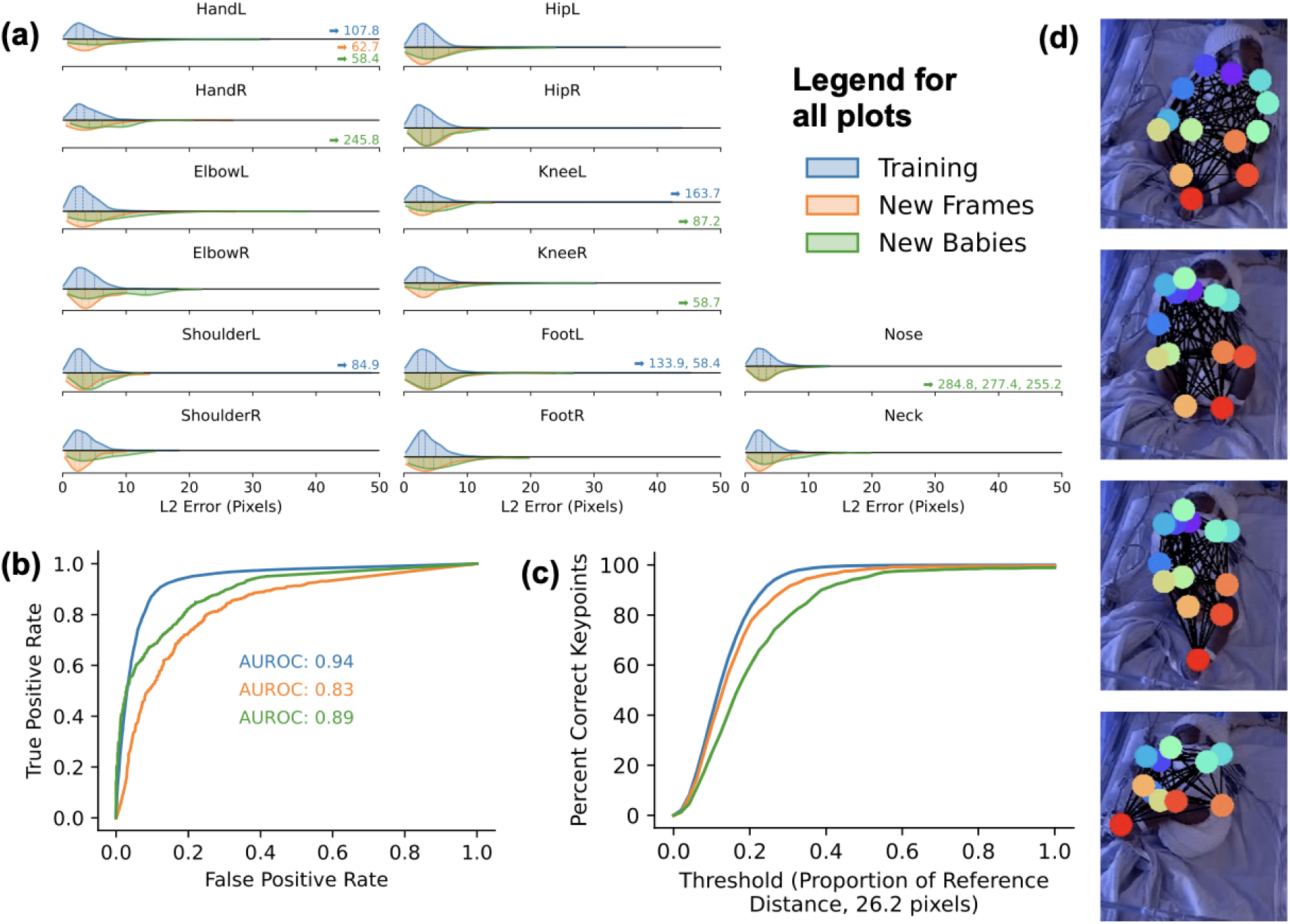
Performance of pose recognition with DeepLabCut on a ResNet-50 backbone. **(a)** The L2 pixel error was comparable to the literature and consistently low across all labeled non-occluded anatomic landmarks for training data (blue, median 3.2 pixels, N=18,399), frames held out from training (orange, median 3.5 pixels, N=797), and frames from infants held out from training (green, median 4.6 pixels, N=973). Thirteen labeled landmarks (0.06%) had a L2 error >50 pixels and their errors are shown with arrows. **(b)** Pose tracking had high area under ROC curves when predicting landmark occlusion. **(c)** The percentage of key points within a reference distance threshold (26.2 pixels, half the height of a head) was >95% for training and test data. **(d** Exemplary video frames from an infant with *KCNQ2* epilepsy syndrome show the predicted anatomic landmarks during a seizure.

### Association between infant movement, predicted by pose AI, and neurologic changes

To evaluate the utility of pose recognition in a typical intensive care unit, we used movement variance as an intuitive metric of infant movement that would reflect major clinical features including PMA, cerebral dysfunction, and use of sedative medications. The proportion of videos with sufficient information to calculate variance per 1-minute intervals for at least 7 body parts was 47% (range 0–100% per infant). Movement increased with corrected age (**Figure S2**) in infants with EEG abnormalities or sedative medications (47-fold increase between lowest and highest age group, permutation P=1x10^-4^, 10,000 permutations) and in those without (15-fold increase between lowest and highest age groups, permutation P=2.4x10^-3^).

We next evaluated if movement decreased in infants receiving sedative medications and in those with cerebral dysfunction. We restricted these hypothesis tests to neonates with PMA <44 weeks because the relationship between age and movement was non-linear and possibly dependent on underlying neuropathology (see **Figure S1a**). We observed decreased movement in infants with cerebral dysfunction (**Figure S1b,** P=1x10^-4^), phenobarbital (P=3.6x10^-3^), both (P=1x10^-4^), and those receiving sedative infusions (P=1x10^-4^). Taken together, we show that movement increased with age and decreased with sedative medications and with cerebral dysfunction, decreasing more so with their combined effects.

### Pose AI predicts sedation and cerebral dysfunction

The movement variance used in the prior section was used to validate pose AI for clinical applicability but does not account for the relative importance of different anatomic landmarks or their relationship. To overcome this limitation, we developed prediction models for sedation and cerebral dysfunction. We further filtered data to stabilize variance estimates and ensure sufficient data per patient (see **Methods** and **Figure S1**), resulting in 118,823 minutes from 63 infants used for model development (range 65–14,409 minutes per infant). XGBoost^33^ had substantially better performance compared to logistic regression and support vector machines across all metrics for sedation prediction (**Table S1**), so it was used for all downstream applications. When predicting sedation (**Figure 3a**, left panel), our classifier had high performance in training data (median ROC-AUC=0.90), held-out minutes (ROC-AUC=0.91), and a set of infants not used for training (AUROC=0.87). We observed similarly high performance with our cerebral dysfunction classifier (**Figure 3a**, right panel) in training data (median AUROC=0.91), held-out minutes (AUROC=0.90), and held-out infants (AUROC=0.76). We interrogated both classifiers for feature importance, and found that feet and shoulders were consistently the most important (**Figure 3b**).

**Figure 3.**
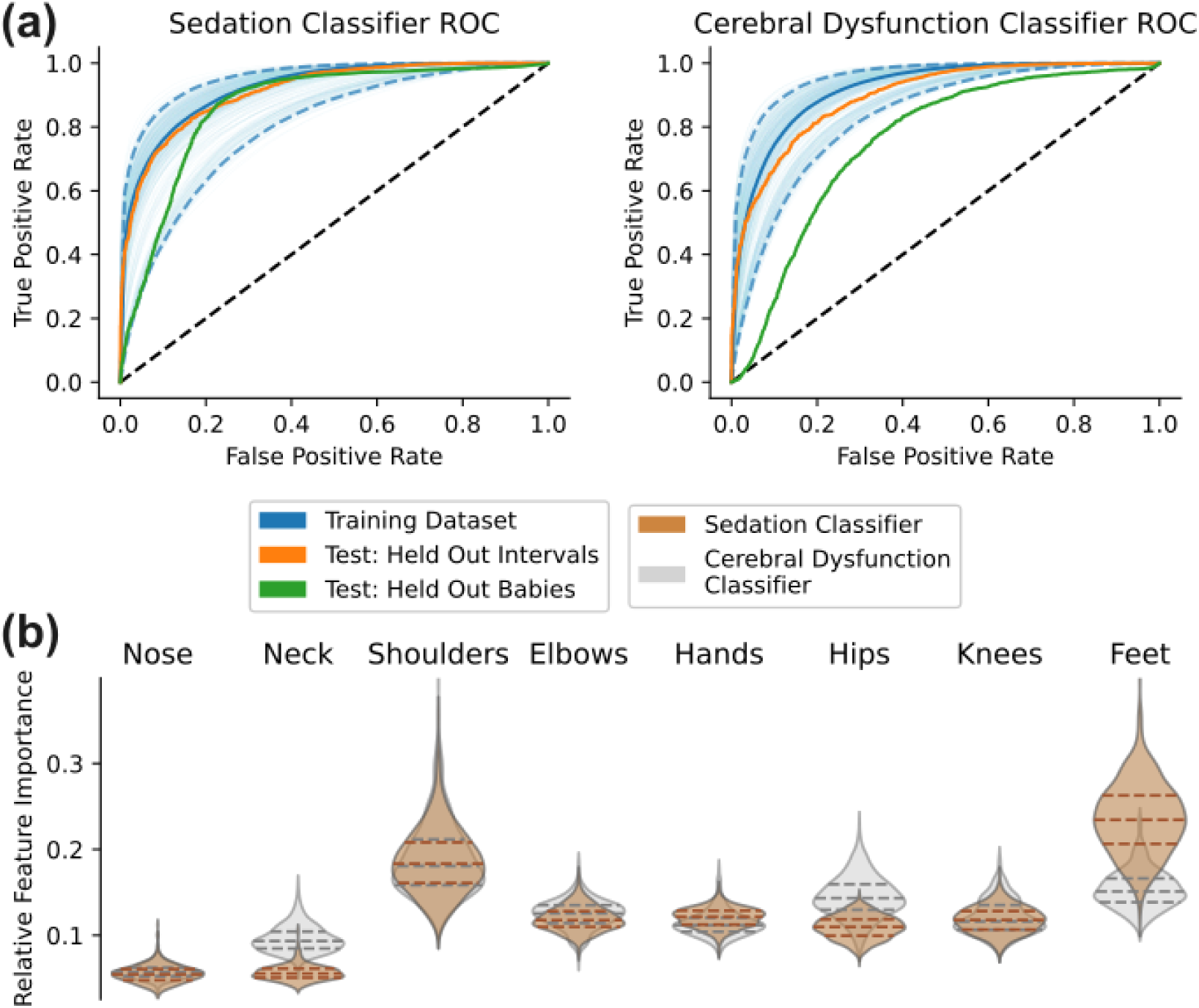
Classifiers trained on infant movement accurately predict sedation and cerebral dysfunction. **(a)** XGBoost receiver operating characteristic curves for sedation (left panel) and cerebral dysfunction classifiers (right panel) trained on pose AI predictions. XGBoost ROC-AUCs are from subsets of the training dataset (blue, median as solid line, 2.5th and 97.5th percentiles as dashed lines), and on test datasets of held-out frames (orange) and held-out infants (green). The dashed black line is a reference, indicating the baseline performance of a classifier operating purely by chance (ROC-AUC=0.5). **(b)** Feature importance from 100 repeats of five-fold cross-validation for both classifiers show that, as expected, anatomic landmarks from the limbs are more important than the nose and neck when predicting sedation and cerebral dysfunction.

### Sedation prediction in an infant undergoing therapeutic hypothermia

To illustrate the utility of predicting sedation in an individual case, we plotted sedation probability as a function of time in an infant born at gestational age 37+1 weeks with Apgar scores 1, 4, and 5, at 1, 5, and 10 minutes, who required therapeutic hypothermia for probable hypoxic ischemic encephalopathy (**Figure 4**). The newborn received phenobarbital before rewarming due to increasing epileptiform discharges observed on EEG. Sedation probability increased first with hypothermia induction and again after phenobarbital administration (P(*Sedation*)_median_ = 0.23 from 1476 minutes before phenobarbital versus 0.59 from 2268 minutes after, Wilcoxon-rank sum test P<10^-10^). Exemplary video frames with heatmaps of infant motion during the preceding five minutes visually capture this decrease in movement (**Figure 4d**).

**Figure 4.**
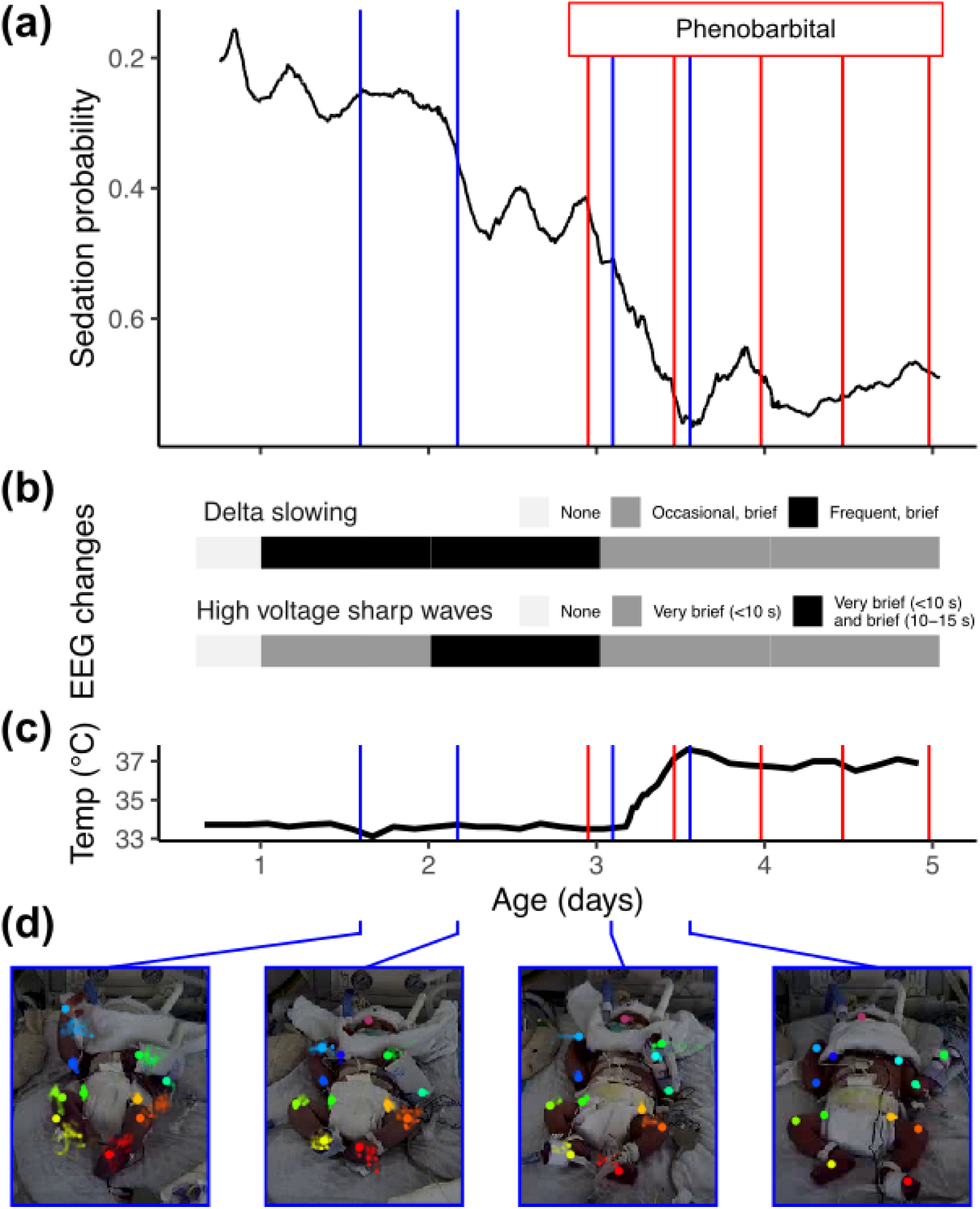
Pose AI predicts sedation during therapeutic hypothermia for an infant with hypoxic ischemic encephalopathy. **(a)** Sedation probability (y-axis) as a function of age in days (x-axis) for a full-term infant transferred to Mount Sinai Hospital for therapeutic hypothermia. Sedation probability increased after hypothermia induction (see panel c) and after phenobarbital administration (red lines). **(b)** The EEG had increased epileptiform discharges at two and three days old, so phenobarbital was administered to protect against seizures during rewarming. **(c)** Temperature during hypothermia and rewarming. **(d)** Exemplary frames during increasing levels of sedation, each annotated with a heat map depicting infant movement over the prior five minutes.

## DISCUSSION

We show that pose AI can accurately track infant movement and predict sedation and cerebral dysfunction in critically ill infants in their “natural” clinical setting. We envision a system for neurologic monitoring akin to cardiorespiratory telemetry. A camera is facing the incubator to record and store video data. Pose AI then provides a neuro-telemetry strip with anatomic landmark position and algorithmic predictions including level of sedation and cerebral dysfunction. If the algorithm detects abnormalities or there is a clinical concern (*e.g.*, tolerance to sedative medications), then the stored video footage, pose tracking, and relevant predictions can be reviewed. This system has high interpretability for bedside providers because reviewing videos is intuitive. Such interpretability is not the case with other infant motion sensing technology (*e.g.*, wearables, mattresses, radar, or waveform artifacts).^35^ Similar workflow integrations are used to monitor for arrhythmias and apnea-bradycardia events.

There is broad applicability of our technology because infants are routinely sedated. Common reasons for sedation are mechanical ventilation, bedside procedures, imaging, therapeutic hypothermia, extracorporeal membrane oxygenation, and clinical pathology such as pulmonary hypertension and unrepaired Tetralogy of Fallot.^5^ Oversedation leads to prolonged mechanical ventilation, brain injury, and drug withdrawal syndromes.^36^ Undersedation is associated with pain, pulmonary hypertensive crises, and adverse events including repeat imaging, catheter displacement, and unplanned extubation.^37^ Pose tracking has potential to address these sedation complications through more precise titration.

Infants are also frequently evaluated for encephalopathy. Cerebral dysfunction is an EEG finding that can reflect encephalopathy, which can be iatrogenic or due to an unknown clinical pathology. For example, it could be due to medications, hypoglycemia, sepsis, hyperammonemia, or permanent injury after neonatal stroke or seizures.^3^ We demonstrate that pose tracking can predict cerebral dysfunction in infants, suggesting utility for identifying and monitoring encephalopathy.

There are multiple strengths in our study. We use explainable features (movement of anatomic landmarks) trained on a large, diverse patient population (25.2% white non-Hispanic, 46% female), addressing important concerns about bias and interpretability with deploying AI.^38^ One previous challenge is that pose AI trained on adults has worse performance on infants,^39^ which we overcame by training on patients less than one year old. Pose AI and neurologic predictions had high accuracy, including on held-out test datasets. This is in contrast to prior studies, which were small (N<30) and did not associate the learned infant pose with neurologic changes in the NICU. One study correlated pose with infant neuromotor risk,^28^ but this study was also small (19 infants, 9.7 hours of video) and placed infants in a bespoke setting outside the NICU. We related variance in movement to neurologic status, analogous to the relationship between heart rate variability and autonomic status.^40^ One limitation of pose AI is infant occlusion due to swaddling, however, 47% of video data was sufficient for movement prediction. Taken together, our results suggest that pose AI monitoring is feasible in the NICU.

Our work describes new methods for neuro-telemetry and their application, and there are important limitations and extensive future directions. The models were trained on data collected at a single institution, and the pose algorithm and neurologic predictions need to be evaluated on video data from other institutions and technologies. This work was also conducted on retrospectively collected video-EEG data. Future adjustments to train more precise models will need to be conducted on prospective NICU cohorts with continuous video data available from birth. For example, in our study, sedation level was based on administration of sedative medications. Prospective work could utilize standardized sedation or analgesia scales.^37^ Cerebral dysfunction is an EEG-derived sign of encephalopathy, but future work can use other markers of encephalopathy like changes in feeding, respiration, or autonomic function. In addition, prospective video studies could correlate infant movement with other clinical changes not typically captured on video EEG, likely sepsis, seizures before loading ASMs, and withdrawal.

Artificial intelligence promises to transform pediatrics but has seen limited applications for one of the most consequential outcomes– neurologic changes in the NICU.^38^ The neurologic assessment has wide-ranging consequences for healthy development, but neuro-telemetry has remained elusive in most NICUs, despite decades of work in EEG and specialized neuro-NICUs. As NICU deregionalization continues,^41^ pose AI may fill this unmet need as a minimally invasive, interpretable, and scalable approach to continuous neuro-monitoring in the NICU.

## METHODS

### Patient population and human subjects’ protection

We collected data through a retrospective observational experimental design between February 01, 2021 and December 31, 2022. This approach was approved by the institutional review board at the Icahn School of Medicine at Mount Sinai. To train algorithms generalizable across race, sex, and gestational age, we used broad inclusion criteria and obtained data from a racially and ethnically diverse patient population. We included data from all infants who had video-EEG data and chronologic age ≤1 year at the start of recording. The Mount Sinai Hospital in New York City is an ideal setting because it serves many newborns (6,000-8,000 births/year), serves a racially diverse population (Asian 6%, Black 13%, White 38%, Other 27%, Unknown 19%), and has broad geographic representation (Manhattan 38%, other Boroughs 37%, and outside NYC 25%), based on demographics for mothers of newborns admitted to the Mount Sinai NICU or well-baby nursery. We transferred all video-EEG and electronic health record data from the Epilepsy Monitoring Unit over an encrypted connection directly to Mount Sinai’s HIPAA-compliant supercomputing cluster, where they were stored. The full data workflow, which follows Transparent Reporting of a multivariable prediction model for Individual Prognosis Or Diagnosis (TRIPOD) guidelines, is shown in **Figure** S1.18,19

### Clinical outcomes

We evaluated the utility and predictive capability of pose AI on two clinical outcomes, sedation and cerebral dysfunction. We classified phenobarbital and sedative infusions (*i.e.*, midazolam, dexmedetomidine and fentanyl) as sedating.^20^ Levetiracetam and less frequently used antiseizure medications (ASMs), such as phenytoin and oxcarbazepine, were not considered sedating for this study.^20^ The observed ASM combinations are enumerated in **Table 1**. Cerebral dysfunction or encephalopathy was diagnosed from EEG readings by an epileptologist separately for each calendar day.

The diagnosis of cerebral dysfunction and encephalopathy was based on focal or generalized abnormalities of background EEG activity, such as reactivity, synchrony, discontinuities, and slowing.

### Software development

There are multiple algorithms to track human pose from video data. Most are trained on adults and have poor performance on infants, likely because infant body proportions are different from adults.^21^ We chose DeepLabCut for its generalizability to many animals,^16,22^ including humans,^23^ and robots.^24^ DeepLabCut uses transfer learning to leverage ResNet,^25^ which is trained on >1 million images, for pose tracking. To train DeepLabCut to recognize infant pose, we randomly sampled 25 frames per infant and labeled up to 14 landmarks per frame. We excluded frames if all body parts were occluded, if the infant was out of frame, or if subsets of the video were incompatible with DeepLabCut, leaving 2712 frames from 109 infants for model training and evaluation. To test generalizability and prevent data leakage,^26,27^ we randomly excluded 10% of infants (N=239 frames, 10 infants) from training and 5% of frames (N =122 frames) from infants used in training. We trained DeepLabCut on 2351 frames from 99 infants and evaluated performance within these training data with repeated measures k-fold cross-validation. With these model weights, any infant video can be converted to vectorized pose, which consists of X and Y coordinates and a likelihood for each anatomic landmark in each frame.

We used three evaluation metrics to assess the performance of the infant DeepLabCut model. First, we generated L2 pixel errors by measuring the Euclidean distance between predicted body part coordinates and their manually labeled anatomic landmarks. Second, we generated area under receiver operating characteristic (AUROC) curves based on the model’s ability to correctly predict whether an anatomic landmark was occluded using DeepLabCut’s built-in confidence score. Finally, we used a standard pose recognition evaluation metric, percentage of correct key points (PCK).^23^ The PCK is the percent of body part coordinates within 26.2 pixels of a landmark’s known position (26.2 pixels is on average half the height of an infant’s head in our video frames).

### Statistical analysis

We first developed a summary metric of infant neurologic status. To quantify baseline movement, we calculated variance for each X and Y position for each body part per minute. We divided variance by median nose-to-neck distance within each interval to adjust for infant size and camera position. Variance was chosen because it summarizes frequency and amplitude of motion while maintaining robustness to outliers. We used variance instead of kinematic calculations, which were used in other infant pose recognition work,^28^ as kinematics were challenging to calculate from two-dimensional videos across the wide variety of camera positions and combination of occluded anatomic landmarks seen in video-EEG data. We also chose variance for its demonstrated clinical relevance with infant mental status: a metric related to variance in infant pose, standard deviation derived from ECG-motion artifacts, was previously shown to predict lethargy in late onset sepsis.^29^ We calculated movement variance within 1-minute intervals per anatomic landmark and then took the median across all landmarks. We then compared 1-minute movement intervals between different groups (e.g., sedation vs no sedation) using a resampling procedure to account for repeated measures (see **Supplementary Information**).^30,31^ A P value less than 0.05 was considered statistically significant.

### Machine Learning

We next used 1-minute movement intervals to develop classifiers for sedation and cerebral dysfunction. Machine learning performance hinges on the quality of input data, so we implemented the following filters (**Figures S1**): postmenstrual age (PMA) <44 weeks to increase homogeneity, ≥7 body parts visible per 1-minute interval to ensure sufficient visibility, variance calculations based on ≥30 seconds per 1-minute interval to ensure stable variance estimates for each anatomic landmark, and ≥60 minutes of usable footage per infant to ensure sufficient sample size per infant. The filtered dataset used for developing sedation and cerebral dysfunction classifiers thus comprised 118,823 minutes from 63 infants (**Figure S1**).

We trained three models to predict sedation, logistic regression, support vector machines, and XGBoost (see **Supplementary Information** for hyperparameter tuning). We selected the best classifier with multiple metrics (F1 score, accuracy, AUROC, precision-recall ROC).^32,33^ Then, we evaluated the best classifier on training data through repeated measures k-fold cross-validation and on two randomly selected test sets, held-out infants (*i.e.*, those not used in training) and held-out minutes from infants used in training. To better understand which anatomic landmarks were contributing to classification performance, we calculated feature importance using the XGBoost ‘gain’ function. To generate a resampled distribution for gain, which is a deterministic function of the XGBoost classifier, we recalculated gain through random subsampling of 80% of the data and repeated this 500 times.

## ACKNOWLEDGEMENTS

We thank the families for entrusting us with the care of their critically ill children. This work was funded by the Friedman Brain Institute Fascitelli Scholar Junior Faculty Grant and Thrasher Research Fund Early Career Award (F.R.). This work was supported in part through the computational and data resources and staff expertise provided by Scientific Computing and Data at the Icahn School of Medicine at Mount Sinai and supported by the Clinical and Translational Science Awards (CTSA) grant UL1TR004419 from the National Center for Advancing Translational Sciences. Research reported in this publication was also supported by the Office of Research Infrastructure of the National Institutes of Health under award number S10OD026880 and S10OD030463. The content is solely the responsibility of the authors and does not necessarily represent the official views of the National Institutes of Health.

## AUTHOR CONTRIBUTIONS

Felix R conceived and designed the study. AG, NB, NA, MLVT, MF, and Felix R acquired, extracted, and/or pre-processed data. AG and Felix R analyzed the data. AG, Florian R, RF, EH, BG, SUM, KG, GNN, and Felix R interpreted the data. AG and Felix R wrote the manuscript. All authors read, revised, and approved the final manuscript.

## DATA AVAILABILITY

The datasets used and/or analyzed during the current study are available from the corresponding author on reasonable request.

## CODE AVAILABILITY

Code is available on github (https://github.com/Agleason1/Neonatal-Pose-AI--Sedation-and-Cerebral-Dysfunction).

