## Supplemental Information for "Accurate prediction of neurologic changes in critically ill infants using pose AI"

Alec Gleason, *et al.*

**Supplemental Methods**

Permutation testing for statistical significance

We calculated the difference in median movement between two groups and compared it to a null distribution.^1,2^ To generate this null distribution, for each movement interval, we first randomly resampled subject metadata (*i.e.*, date of study, current medications, and EEG abnormalities) with replacement. We then calculated the difference in median movement between two groups (*e.g.*, sedation vs no sedation) using the randomly resampled data. We repeated this resampling procedure 10,000 times to generate a null distribution of differences. The P value was the proportion of results as or more extreme than observed.

XGBoost fine-tuning

We fine-tuned XGBoost^3^ hyperparameters via 5,000 simulations of Random Search and identified the best hyperparameters through repeated k-fold cross-validation. For each iteration of Random Search, the values for 12 hyperparameters were randomly assigned from a prespecified range and performance is calculated as the mean F1 score from three repeats of five-fold cross-validation. We used a macro-averaged F1 score to account for potential class imbalances.

**Supplemental Figures**

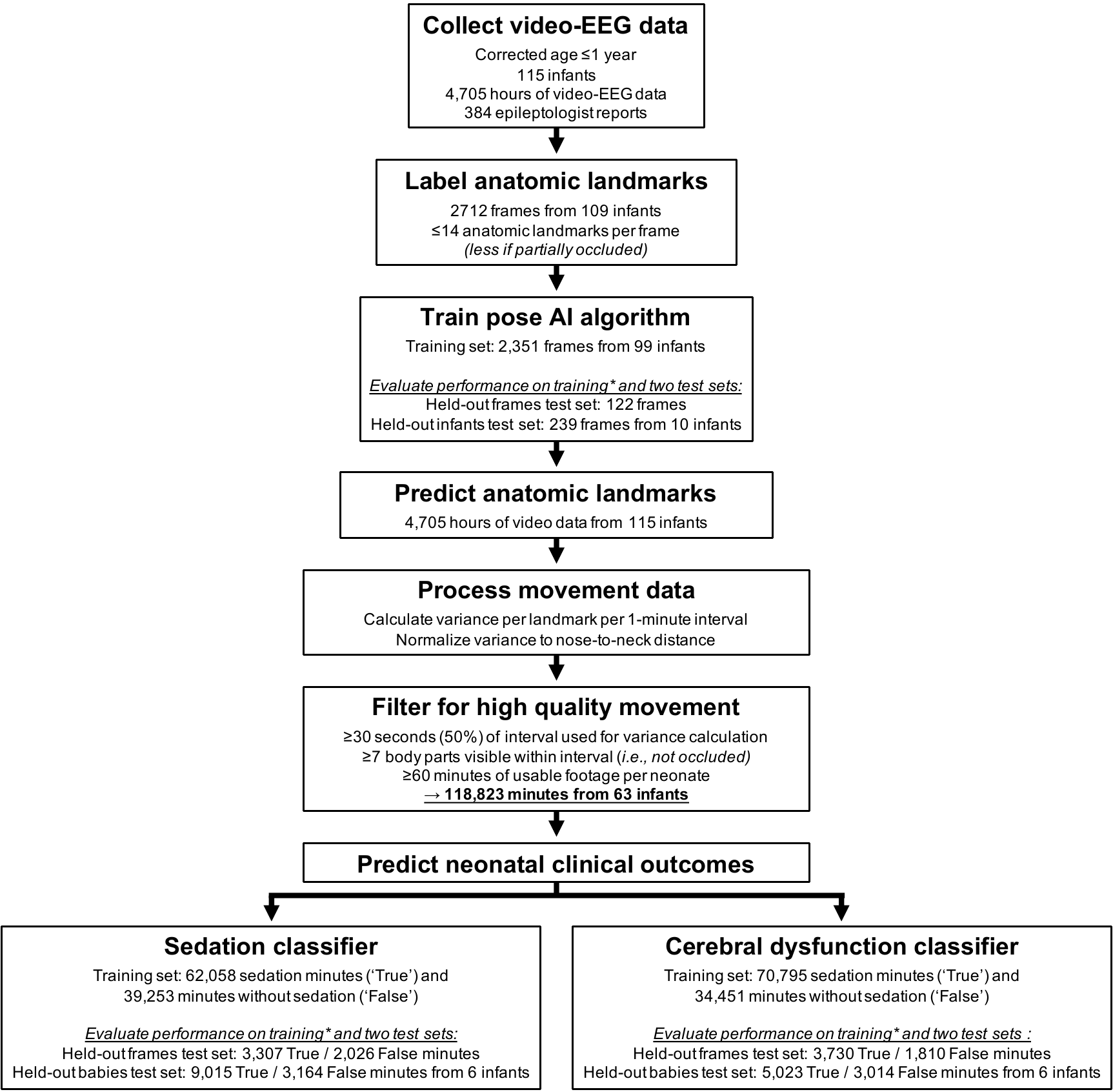

**Figure S1. Data filtering workflow.** The full data workflow including filters, sample sizes for each group, and train / test datasets, comporting with TRIPOD guidelines.^4,5^ *Performance was evaluated on training data through k-fold repeated measures cross-validation. Performance was also evaluated on two held-out test sets, held-out infants and held-out frames from infants used in training.

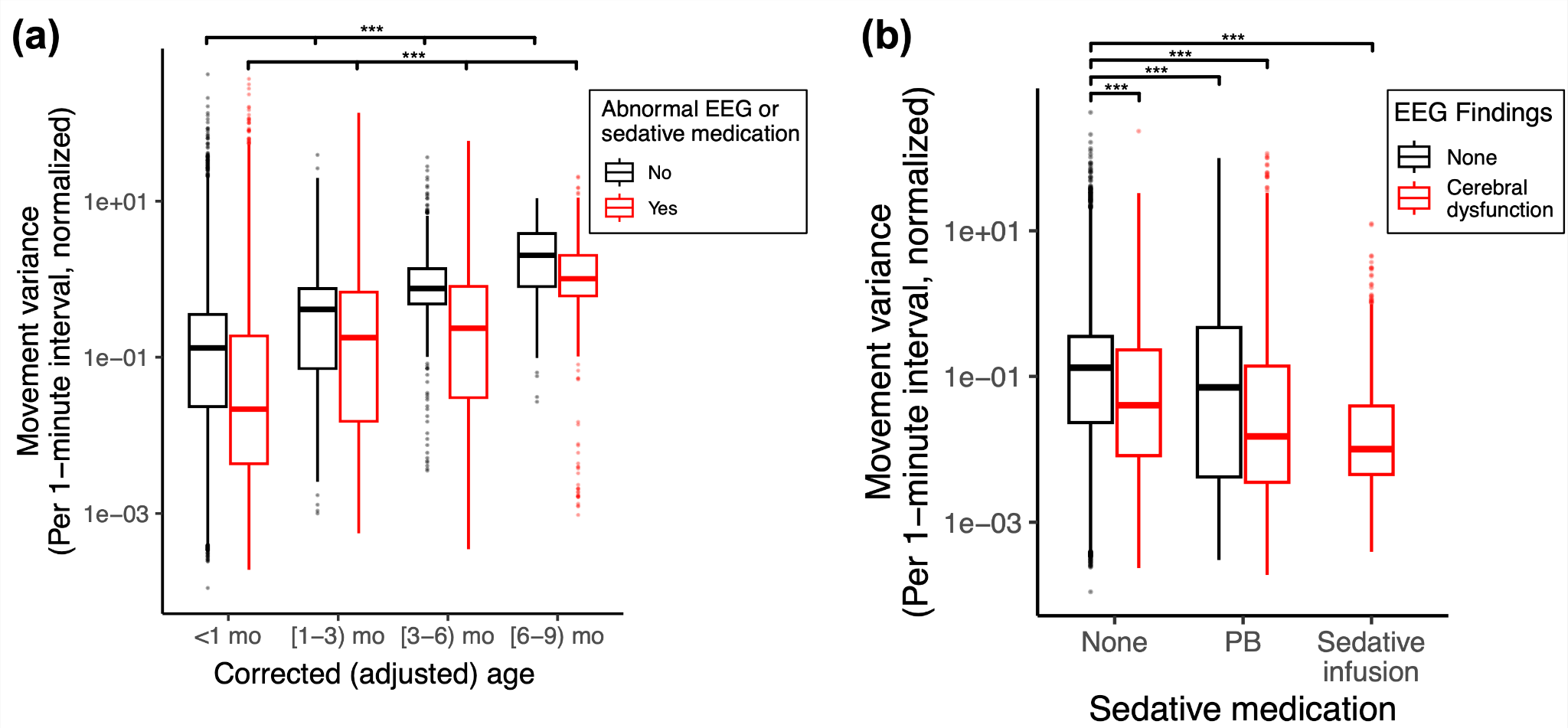

**Figure S2. Infant movement, predicted using pose AI, increased with age and decreased with sedative medications and EEG abnormalities.** **(a)** Movement variance within 1-minute intervals (y-axis, log-scaled) increased with corrected age (x-axis) in patients with sedative medications or abnormal EEGs (red, N=110,622 minutes, 53 infants) and in those without (black, N=29,093 minutes, 46 infants). **(b)** Among infants <44 weeks postmenstrual age, movement was lower with cerebral dysfunction (N=21,541 minutes, 21 infants), phenobarbital (N=1,488 minutes, 3 infants), both (N=55,724 minutes, 24 infants), or those receiving sedative infusions (N=5378 minutes, 3 infants) when compared to infants with normal EEG and neither medication group (N=27,099 minutes, 35 infants). This was also statistically significant for all permutation tests (a permutation *P*-value of <10^-3^ is labeled with ***). PB=phenobarbital.

**Supplemental Tables**

**Table S1.** Performance of sedation classifiers on training data obtained via five-fold cross-validation.

| **Model** | **ROC-AUC** | **AUPRC** | **F1 Macro Score** | **Accuracy** |
| --- | --- | --- | --- | --- |
| Logistic | 0.64 | 0.71 | 0.60 | 0.65 |
| SVM | 0.68 | 0.74 | 0.58 | 0.66 |
| XGBoost | **0.94** | **0.96** | **0.86** | **0.86** |
